# Effects of childhood adversity on socially learned placebo analgesia in virtual reality: A cross-sectional study

**DOI:** 10.64898/2025.12.04.25341572

**Authors:** Lakota Watson, Yang Wang, Jewel N White, Roni Shafir, Giancarlo Colloca, Jonathan Michael Heagerty, Sida Li, Barbara Brawn, Amitabh Varshney, Shuo Chen, Luana Colloca

## Abstract

Early life adversity (ELA), indexed through Adverse Childhood Experiences (ACEs), is associated with long-term alterations in emotion regulation, stress responsivity, and social learning—factors that may shape learned pain modulation. Social observation in immersive virtual environments offers a powerful way to investigate these mechanisms.

To examine whether ELA influences social observation-induced placebo analgesia and empathy responses in immersive and non-immersive contexts.

Adults with self-reported low versus high ACE exposure completed an observational learning task in immersive virtual reality (VR) or a non-immersive format. Participants observed Human or Avatar demonstrators experiencing pain relief and then underwent self-pain testing. Measures included socially induced placebo analgesia and affective and cognitive components of state empathy.

Individuals with high ACE exposure showed stronger social observation-induced placebo analgesia, particularly within immersive VR. High ACE participants exhibited reduced affective state empathy, while cognitive empathy remained comparable to the low ACE group.

Elevated ELA is unexpectedly associated with enhanced responsiveness to socially learned placebo analgesia, especially in immersive VR settings. These findings highlight how early adversity may shape sensitivity to socially transmitted treatment cues, with implications for the design of VR-based therapeutic interventions.

## Introduction

Early life adversity (ELA) [14], commonly indexed by Adverse Childhood Experiences (ACEs) [14; 28], is associated with enduring alterations in emotion regulation, stress responsivity, and social learning [19; 35; 41; 52; 62]. Individuals with high ACE exposure often show dysregulated hypothalamic–pituitary–adrenal (HPA) axis function and reduced prefrontal control [15; 24; 26; 59; 63], which is hypothesized to increase vulnerability to chronic pain and mood disorders [22; 27; 29; 40; 50]. Beyond influencing emotional and physiological reactivity, these alterations may also affect how individuals learn from and adapt to new environments, particularly those that rely on social and contextual cues to shape behavior [11; 16]. Understanding how ELA modulates learning-based pain regulation in digital health can therefore provide insight into mechanisms that underline individual differences in intervention responsiveness.

Observational learning where individuals acquire expectations of pain relief by observing others [43] offers a framework for studying learning-based modulation of pain [3; 4; 10; 32; 39]. These procedures engage social and cognitive systems [38; 42] that are often altered by early adversity [37; 41], making them suitable for investigating how ELA modulates pain-related learning. Integrating these social learning procedures with immersive technologies [48; 58] provides a way to bridge laboratory models of social learning with ecologically valid contexts that more closely mirror real-world experiences.

Immersive virtual reality (VR) offers such a platform. By combining sensory and social realism with experimental control, VR enables systematic manipulation of contextual and interpersonal cues while measuring behavioral, physiological, and neural data [23; 36]. This makes VR uniquely suited to study mechanisms through which state empathy and social learning modulate pain responses. However, despite the increasing use of VR in pain management and rehabilitation, no studies to our knowledge have examined how developmental histories such as ACE exposure influence placebo responsiveness in immersive digital environments.

We recently extended an observational learning procedure [10; 39] into immersive VR to examine socially induced placebo analgesia. Participants observed Human or Avatar demonstrators experiencing pain relief while their behavioral and physiological responses were recorded. We found that placebo effects are larger in VR when paired with an Avatar demonstrator than a Human demonstrator while in the non-VR context, a Human demonstrator induces larger placebo effects [56].

We hypothesized that VR would amplify observationally learned placebo effects by enhancing the salience and realism of social cues, and that these effects would vary as a function of exposure to ELA. To test these hypotheses, we implemented a previously published social observational learning method in immersive VR^33^ stratifying participants by ACE. Specifically, we expected that individuals with higher ACE scores would show reduced social observation–induced placebo effects and lower affective state empathy for pain.

## Methods

### Study design

This cross-sectional observational study examined how exposure to adverse experiences in childhood influences socially induced placebo analgesia and state empathy in immersive and 2D environments using thermal pain stimulation. This study is an extension of a previous project whose main findings have been reported elsewhere [56]. Building on that work, the present study specifically investigates how exposure to ACEs modulates socially induced placebo analgesia in digital and immersive environments.

### Recruitments

Healthy participants were recruited from a university campus, via social media advertisements, and through Build Clinics, for a two-day within-subjects experiment conducted at the University of Maryland, Baltimore School of Nursing between May 2022 and September 2023. Participants who consented to additional data collection completed a brief questionnaire assessing exposure to adverse experiences in childhood and adolescence. The final analytic sample included 36 adults (age, *M* = 31 years, range 18–61) who provided both experimental and childhood adversity data. Among them, 25 identified as female, and 17 identified as white (see **Table 1**).

**Table 1.** Demographic characteristics of the participants.

### Ethical considerations

All experimental procedures were approved by the University of Maryland, Baltimore Institutional Review Board (Protocol HP-00085382) and conformed to the Declaration of Helsinki. Prior to entry to the study, researchers explained study procedures and risks, as well as the use of deception, and participants were given the opportunity to ask questions. Participants then provided written informed consent and received compensation of $180 for completing both sessions, with an additional $25 for completing the adversity questionnaire. At study completion, all participants were debriefed regarding the true nature of the observational learning task and given the opportunity to withdraw. No participant withdrew consent. The authors have obtained written consent to publish the images and have applied all reasonable measures to protect demonstrator anonymity.

### Eligibility and Screening

Eligibility criteria included good general health and no history of chronic pain, neurological, cardiovascular, pulmonary, renal or hepatic disease, cancer, psychiatric disorders, or pregnancy. Participants with sensory impairments, VR-induced claustrophobia, or prior adverse VR experiences were excluded. Urine toxicology screens were conducted at each session to rule out amphetamines, cocaine, opioids, or THC. Participants testing positive were excluded from participation.

### Study Procedures

This study investigated how exposure to ELA modulates socially induced placebo analgesia using VR and 2D observation procedures. Participants completed all experimental conditions in a within-subjects design, allowing direct comparison of individual responses across immersive and non-immersive (2D) contexts.

The experiment was conducted over two sessions, scheduled no more than ten days apart, in a controlled laboratory environment at the University of Maryland, Baltimore School of Nursing. On Day 1, participants completed baseline assessments including height, weight, blood pressure, heart rate, and quantitative sensory testing[18] to determine individualized thermal pain thresholds. Pain calibration used brief heat pulses to the left ventral forearm, ensuring subsequent experimental stimuli were perceived as moderately painful (VAS 50–60). Participants also completed screening questionnaires and the Implicit Association Task[20] to account for potential biases.

Day 2 focused on experimental manipulation. Participants completed four counterbalanced conditions: Human VR, Avatar VR, Human 2D, and Avatar 2D. Each condition consisted of two phases: an observation phase, in which participants viewed a demonstrator (Human or Avatar) receiving thermal stimulation paired with visually distinct creams (blue or green, both inert), and a self-experience phase, in which participants received identical thermal stimuli to the same left ventral forearm site. Observation videos modeled placebo effects by showing lower pain ratings for one cream (10–30 VAS) relative to the other (70–90 VAS).

During the observation phase, participants attended visual cues and video sequences and subsequently rated their state empathy on affective (self-referred) and cognitive (other-referred) dimensions. The VR environment was implemented via a head-mounted display with interactive VAS tools, allowing participants to provide ratings in real time. This digital platform enabled precise control of visual perspective, demonstrator identity, and environmental context while capturing continuous behavioral metrics such as response latency and engagement.

In the self-experience phase, participants received thermal stimuli cued by the same color-coded creams. They rated perceived pain intensity and unpleasantness using the VR-integrated VAS. Placebo analgesia was calculated as the difference between control and “treatment” trials on a trial-by-trial basis. Details are shown in **Figure 1**.

**Figure 1.**
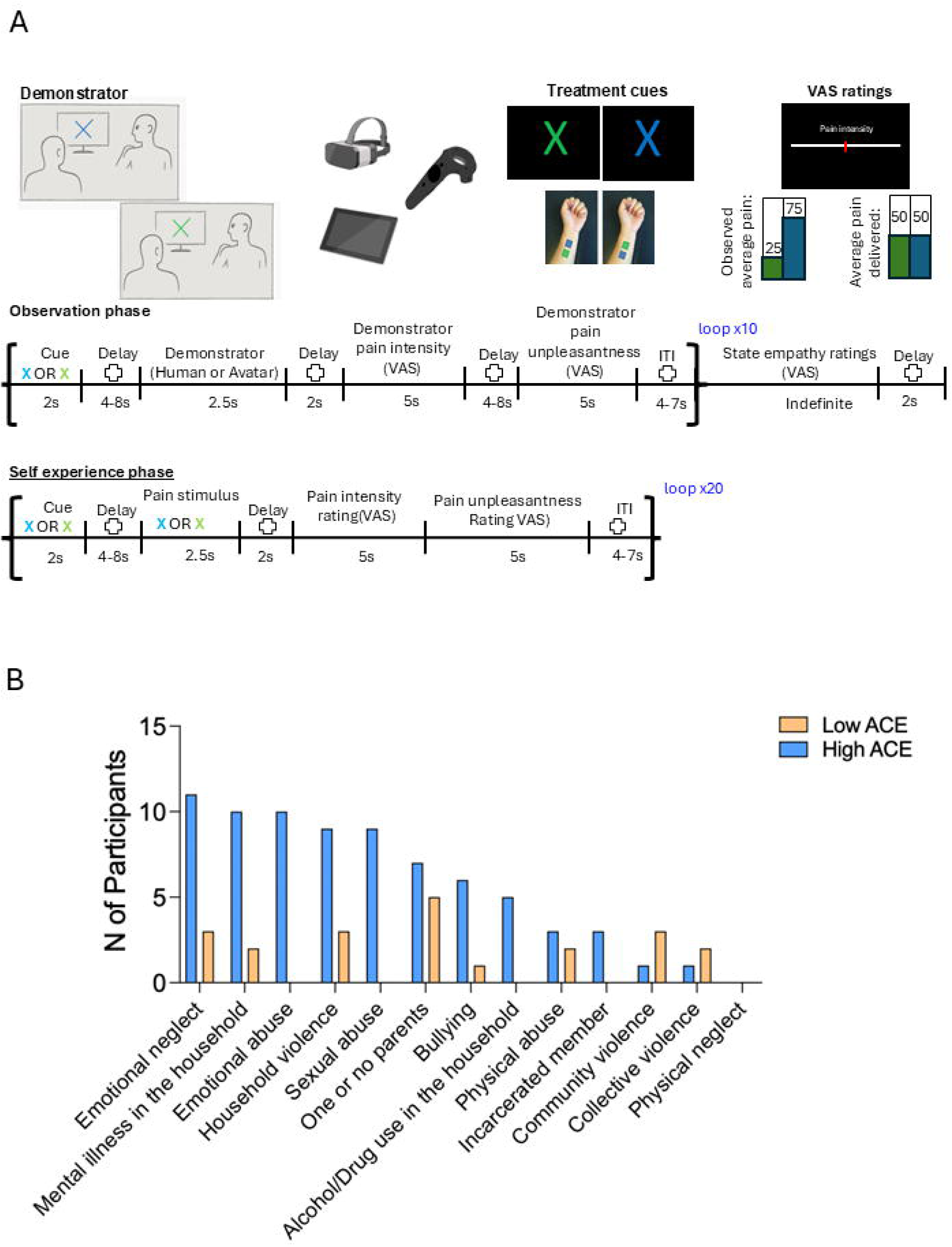
Study design and ACE distribution (B). **(A).** Participants observed Human or Avatar demonstrators receiving heat stimulation on the left forearm following color cues (blue or green) that predicted low or moderate pain intensity (25 vs. 75 on a 0–100 VAS). Demonstrations were presented via immersive VR or 2D tablet video. Participants then rated affective (self) and cognitive (other) empathy. In the subsequent self-experience phase, participants received identical cues before heat stimulation (VAS 50) and rated pain intensity and unpleasantness. Trials were repeated 20 times per cue, and VAS responses were collected via the VR controller. Treatment location was randomized across participants and conditions. ITI = inter-trial interval. **(B)** Distribution of reported ACE categories across participants, showing relative prevalence of emotional neglect, absence of parents, household violence, mental illness in the household, and emotional abuse.

VR implementation leveraged standardized Avatar modeling and immersive laboratory replication to maximize ecological validity and reproducibility. The Avatar was modeled after the Human demonstrator **Figure 1**. By integrating digital interventions, controlled observation learning, and ACE stratification, this design allowed us to examine how early adversity shapes behavioral and neural responses in a scalable, digitally mediated experimental framework, highlighting mechanisms relevant for personalized digital health interventions.

### Measurements

Pain ratings were collected using anchored visual analog scales (VAS, 0–100) for intensity and unpleasantness across all conditions. In VR, participants used joystick-controlled VAS tools integrated within the headset, enabling real-time behavioral data collection. Socially induced placebo analgesia was quantified as the trial-by-trial difference between control and treatment conditions (VAS control – VAS treatment) during the self-experience phase.

Affective state empathy items measured participants’ own emotional and sensory responses to the demonstrator’s pain, whereas cognitive state empathy items assessed participants’ perception of the demonstrator’s experience. Responses were recorded on the same 0–100 VAS.

State empathy measures were obtained at the end of each *observation* condition by asking participants to evaluate the following: a) the intensity of the sensation experienced by the participant themselves or supposedly felt by the demonstrator (sensory; self-referred and other-referred, respectively) and b) the unpleasantness of the sensation experienced by the participant or felt by the demonstrator (emotional; self-referred and other referred, respectively) as per Betti et al. 2009 [7].

Exposure to early life adversity was assessed using the Adverse Childhood Experiences—International Questionnaire (ACE-IQ[57]), covering 14 categories including abuse, neglect, household dysfunction, peer violence, and community or collective violence. Responses were scored frequency-based and participants were stratified into low ACE (0–2 ACEs) and high ACE (≥3 ACEs) groups for analysis. This allowed evaluation of how ELA modulates socially induced placebo responses and state empathy, providing insights into individual variability in digital intervention efficacy.

Trait psychological measures were collected at baseline via validated questionnaires. Trait empathy was assessed by the Interpersonal Reactivity Index (IRI) [12], a 28-item self-report questionnaire that consists of four subscales: a) empathic concern (EC, which assesses the tendency to experience feelings of sympathy and compassion for others in need), b) personal distress (PD, which assesses the extent to which an individual feels distress as a result of witnessing another’s emotional distress), c) perspective taking (PT, which assesses the dispositional tendency of an individual to adopt the perspective of another), and d) fantasy scale (FS, which assesses an individual’s propensity to become imaginatively involved with fictional characters and situations). We also assessed Basic Empathy Scale (BES[25]) assessed general cognitive and affective empathy and Harm Avoidance and Reward Dependence using the Tridimensional Personality Questionnaire [9].

Given the associations between ACE, anxiety, depression, and pain, participants completed the State-Trait Anxiety Inventory (STAI [47]), Beck Anxiety Inventory (BAI [6]), and Beck Depression Inventory (BDI [5]) to account for potential mediating effects. All questionnaires were administered through a secure, privacy-compliant platform (REDCap [21]).

### Data analysis

Data normality was examined using Shapiro-Wilk tests. Shapiro–Wilk tests revealed that the distributions of cognitive state empathy, affective state empathy, and pain ratings were non-normal and positively skewed. Because linear mixed models assume normality of residuals rather than dependent variables, residuals were examined post hoc and found to be approximately normal, with only minor deviations in the tails. Outliers were identified using Tukey’s method with a 2.2 interquartile range (IQR) multiplier, where the upper and lower limits were defined as Q3 + 2.2 × IQR and Q1 – 2.2 × IQR, respectively. Q1 and Q3 denote the 25th and 75th percentiles.

We planned a linear mixed-effects model with 20 repeated trials for each experimental condition. Based on prior literature examining ACE and descending pain modulation,[51] we assumed an effect size of ACE on placebo effects of *r* = 0.204, corresponding to Cohen’s *f* = 0.208. Power was calculated for the main effect of ACE on repeated measures of placebo effects across conditions. Under these assumptions (α = 0.05, 1-β = 0.80), a minimum of 28 participants (14 per group) were estimated to provide adequate power to detect a medium-sized effect.

We used linear mixed-effects models (LMM) to account for repeated measures and within-subject variability. We first determined whether social observation could have induced significant placebo effects in this study. To achieve this, LMM was performed including ACE group (low vs. high), context (VR vs. 2D), demonstrators (Human vs. Avatar), and cream type (placebo vs. control) as fixed factors. Participant-specific intercepts are modeled as random effects. Sex, race, trial order, as well as the color of placebo cream were included as covariates.

Upon confirming the main effects of the cream type (placebo vs. control), we operationalized the primary outcome, socially induced placebo analgesia, as the difference in pain ratings between control and treatment trials (VAS control – VAS treatment) during the self-experience phase. To examine how ACE would have influenced the magnitude of socially induced placebo effects in the experimental conditions, separate LMMs were conducted with ACE group (low vs. high), context (VR vs. 2D), and demonstrators (Human vs. Avatar) as fixed factors with similar random effects and covariates. Interaction terms between ACE group and observation conditions allowed assessment of whether VR environments enhanced or attenuated placebo effects differentially in participants with higher ELA.

State empathy ratings from the observation phase were analyzed similarly to evaluate affective and cognitive components as potential moderators of placebo responsiveness. Similar LMMs were performed on the affective/cognitive state empathy (VAS control – VAS placebo) with ACE group (high vs. low), context (VR vs. 2D), and demonstrators (Human vs. Avatar) as the fixed factors.

We further examined whether depression or anxiety levels mediated the relationship between ACE exposure and empathy ratings induced by the VR and 2D Avatar conditions. Specifically, in the mediation models, ACE exposure (high vs. low) was included as the independent variable (X), depression and anxiety levels as the mediator (M), and state empathy ratings for pain intensity and pain unpleasantness as the dependent variables (Y), separately.

Secondary analyses controlled for baseline anxiety and depression scores to rule out potential confounding effects on pain perception and state empathy. Differences in baseline anxiety and depression and other potential mediators between groups were evaluated using Mann-Whitney U tests (when the dependent variables were not normally distributed) with independent t-test applying Bonferroni corrections for multiple comparisons (when the dependent variables were normally distributed).

All analyses were conducted in SPSS (v.21), with α = 0.05 used to determine statistical significance. Power analysis was conducted using G*Power software (v.3.1.9.6) [13].

## Results

The socio-demographic characteristics of the sample are summarized in **Table 1**. In our cohort, the most prevalent ACE categories were emotional neglect (*n* = 13), single or absent parent (*n* = 12), and household violence (*n* = 12); followed by mental illness in the household (*n* = 11) and emotional abuse (*n* = 9). Participants who reported three or more ACE categories were classified as the High ACE group (*n* = 17), while those with fewer than three ACE categories were classified as Low ACE (*n* = 19). There were no significant differences in age (*t*_34_ = −0.56, *p* = .581), race (*χ²_34_* = 0.14, *p* = .709), or sex (*χ²_34_* = 0.02, *p* = .888) between the High and Low ACE groups.

### Placebo analgesia for pain intensity and unpleasantness

We first tested whether the experimental manipulation effectively induced observationally learned placebo analgesia and whether this effect varied by early life adversity (ACE), immersive context (VR vs 2D), and demonstrator type (Human vs Avatar).

A significant main effect of placebo condition confirmed observationally induced analgesia across contexts for pain intensity ratings, *F*_1, 6527.76_ = 75.10, *p* < .001, Cohen’s *d* = 0.14. Importantly, this effect was modulated by a four-way interaction between ACE group, context, demonstrator, and placebo condition, *F*_1, 4754.46_ = 11.54, *p* < .001, indicating that the magnitude and generalization of learned analgesia varied systematically across conditions.

Post hoc comparisons with Bonferroni correction revealed distinct patterns between ACE groups. Participants with low ACE exposure exhibited significant placebo analgesia only in the 2D Human condition (*M* = 10.56, SEM = 1.51, *p* < .001), but not in VR contexts (VR Human: *p* = .428; VR Avatar: *p* = .216) or 2D Avatar (*p* = .831). In contrast, those with high ACE exposure showed a generalized placebo effect across all conditions (all *ps* < .023), with stronger effects in VR than 2D environments (*p* < .01).

To further quantify these differences, we examined placebo magnitude (ΔVAS = control – placebo) in linear mixed-effects models including ACE group, context, and demonstrator as fixed factors and sex, race, color and location of placebo cream, as well as the experimental sequence as covariates. A main effect of ACE confirmed that High ACE participants exhibited overall stronger placebo effects than Low ACE participants, *F*_1, 2185.35_ = 11.88, *p* < .001, Cohen’s *d* = 0.16. An ACE × context interaction was also significant, *F*_1, 2166.42_ = 39.24, *p* < .001, showing opposite patterns across environments: 1. For Low ACE participants, placebo analgesia was stronger in 2D than VR (*p* < .001). 2. For High ACE participants, placebo analgesia was stronger in VR than 2D (*p* < .01).

Additionally, an ACE × context × demonstrator interaction emerged, indicating distinct social cue processing effects across groups. Among Low ACE individuals, placebo effects were larger when observing Avatars in VR (*p* < .006) and Humans in 2D (*p* < .001). Conversely, for High ACE participants, demonstrator type (Human vs Avatar) did not significantly modulate placebo responses in either context (VR: *p* = .108; 2D: *p* = .818). Individual data are shown in **Figure 2A**.

**Figure 2.**
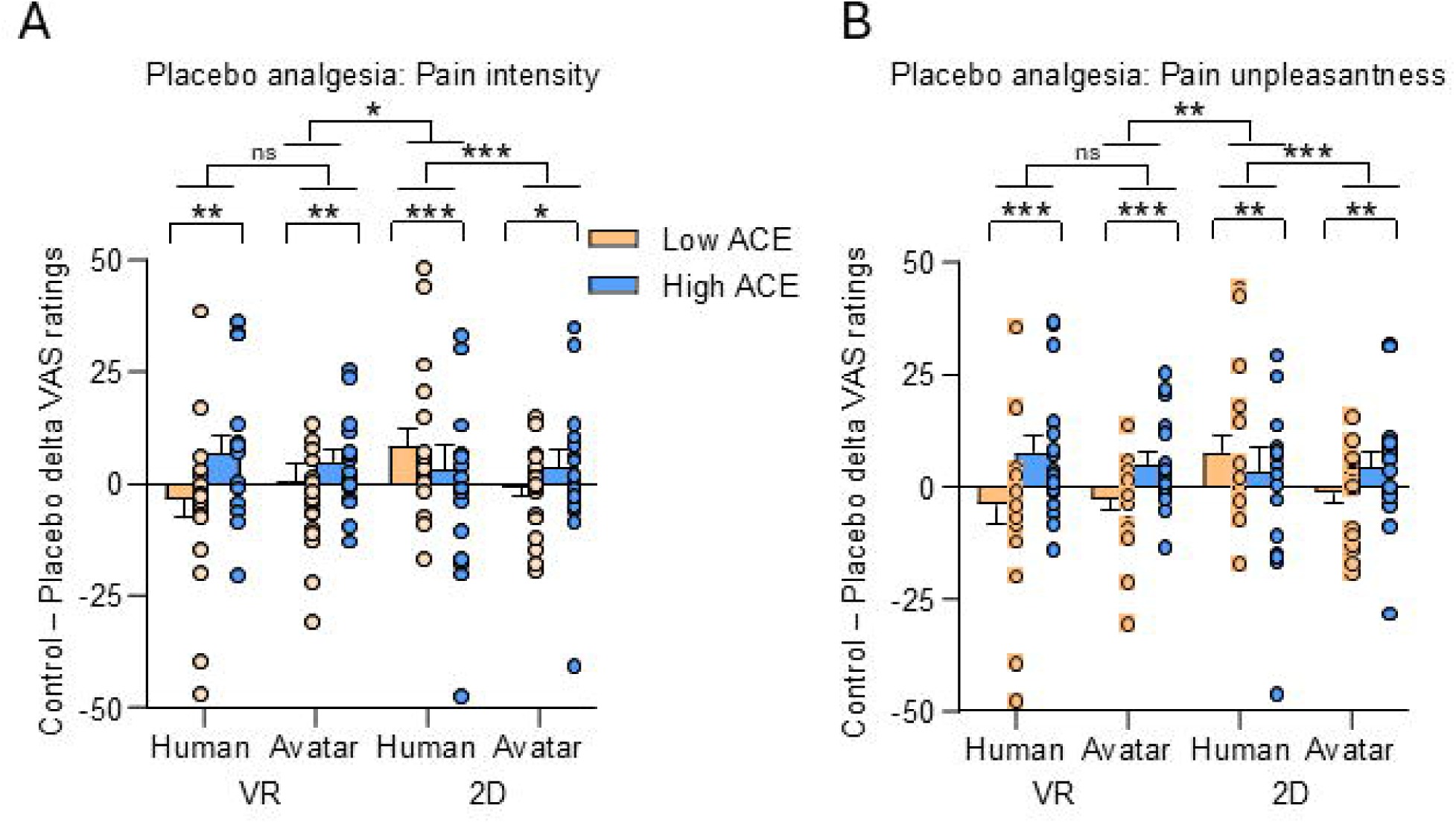
Placebo hypoalgesia for pain intensity (A) and unpleasantness (B) by ACE exposure and context. **(A)** High ACE participants exhibited stronger placebo analgesia, particularly in VR, whereas Low ACE participants showed effects mainly in the 2D Human condition. **(B)** Similar patterns emerged for pain unpleasantness. These findings suggest that early adversity enhances socially learned pain modulation in immersive environments. Group means (±SEM) and individual data points are shown for Low ACE (orange) and High ACE (blue) participants across Virtual Reality (VR) and 2D Human or Avatar demonstrations.

A similar pattern emerged for pain unpleasantness ratings. There was a significant main effect of placebo condition, *F*_1, 6378.20_ = 73.05, *p* < .001, qualified by a higher-order interaction with ACE group, VR/2D context, and demonstrator type (*F*_1, 4619.61_ = 10.07, *p* = .002; see **Figure 2B**).

Among participants with low ACE exposure, the strongest placebo effects occurred following Human demonstrators in 2D *(M=* 9.80, SEM = 1.50, *p* < .001), whereas VR conditions elicited nonsignificant responses (VR Human: *M*= –2.01, SEM = 1.54, *p* = .191; VR Avatar: *M* = –2.18, SEM = 1.15, *p* = .057).

In contrast, participants with high ACE exposure showed robust placebo responses across all contexts (*all ps* < .012), with the largest reductions in pain unpleasantness observed in VR-Human (*M* = 9.24, SEM = 1.62, *p* < .001) and VR-Avatar (*M* = 5.50, SEM = 1.21, *p* < .001) conditions compared to 2D (*p* = .002).

The magnitude of placebo effects differed significantly by ACE exposure, *F*_1, 2267.33_ = 34.08, *p* < .001, with greater analgesia in High ACE participants. Both the ACE × context (*F*_1, 2229.74_ = 55.36, *p* < .001) and ACE × context × demonstrator (*F*_1, 2229.74_ = 22.57, *p* < .001) interactions were significant.

Post-hoc comparisons indicated that, for the Low ACE group, VR-Human and VR-Avatar induced comparable placebo effects (*p* = .212), while 2D-Human produced larger effects than 2D-Avatar (*p* < .001). Conversely, in the High ACE group, VR-Human induced stronger placebo effects than VR-Avatar (*p* = .042), with no difference between 2D-Human and 2D-Avatar (*p* = .864).

Secondary data analysis controlling for baseline anxiety and depression levels revealed similar patterns for both placebo effects for pain intensity (ACE main effect: *p* < .001; ACE × context: *p* < .001; ACE × context × demonstrator: *p* < .001) and pain unpleasantness (ACE main effect: *p* < .001; ACE × context: *p* < .001; ACE × context × demonstrator: *p* < .001).

### State empathy modulation

Using LMMs with ACE group, context, demonstrator, and treatment condition as fixed factors, we found a significant main effect of placebo condition on affective empathy ratings, *F*_1,79.86_ = 13.89, *p* < .001. Observing pain relief (placebo condition) elicited significantly lower affective state empathy for pain intensity (*M* = 1.25, SEM = .23) compared to control observations (*M* = 6.95, SEM = 1.51, *p* < .001), suggesting that witnessing relief cues attenuates emotional resonance with others’ pain.

We computed affective state empathy (VAS_control – VAS_placebo) and examined ACE, context, and demonstrator effects. A significant main effect of ACE group indicated that Low ACE participants reported greater affective state empathy during observation than those with High ACE participants, *F*_1,111.73_ = 8.73, *p* = .004. Among Low ACE participants, the Avatar elicited greater delta affective state empathy responses compared to the Human demonstrator when observed in VR (*p* = .016), **Figure 3A**.

In contrast, high ACE participants displayed blunted affective state empathy responses across all conditions, *F*_1,111.73_ = 8.73, *p* = ns.

**Figure 3.**
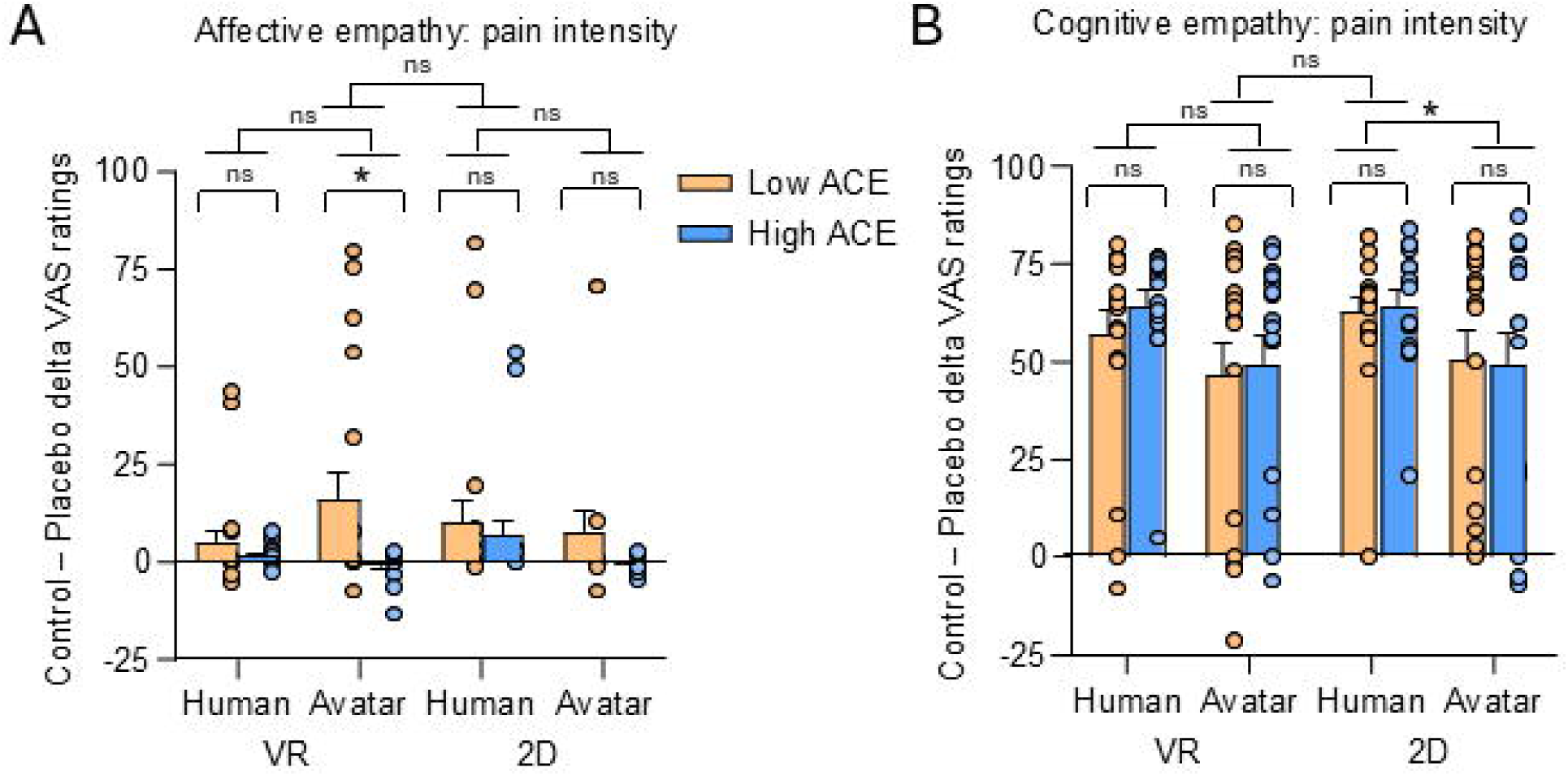
Affective and cognitive empathy modulation by ACE exposure and observation context. **A**. Low ACE participants reported greater affective empathy, especially during Avatar observation in VR, whereas High ACE participants showed blunted responses across conditions. Group means (±SEM) and individual data points are shown for Low ACE (orange) and High ACE (blue) participants during observation of Human and Avatar demonstrators in VR and 2D. **B.** Cognitive empathy did not differ by ACE exposure but was higher during Human compared to Avatar observation. Early adversity thus selectively dampens affective, but not cognitive, empathy in immersive contexts.

Cognitive state empathy did not significantly differ across ACE, *F*_1,112.81_ = 0.41, *p* = .524, though higher VAS scores during Human observation were noted for all participants in comparison with Avatar observation, *F*_1,109.42_ = 8.59, *p* = .004, **Figure 3B**.

We further examined whether individual differences in empathic resonance predict placebo effects within each ACE group. We performed an LMM with delta affective/cognitive state empathy × ACE group × context × demonstrator as fixed factors and magnitudes of placebo effects as the dependent variable. A significant four-way interaction emerged between ACE group, context (VR vs. 2D), demonstrator type (Human vs. Avatar), and delta affective state empathy, *F*_1, 77.642_ = 4.25, *p* = .043, indicating that the magnitude of observationally induced placebo analgesia differed across these factors. Specifically, the pattern of affective the state empathy-placebo effects relationship was context-dependent and varied by both ACE group and demonstrator type, suggesting that immersive social cues modulate analgesic learning differently depending on early life experiences of adversity. No such effects were observed for delta cognitive state empathy scores, *F*_1, 75.23_ = 1.88, *p* = .175.

### Early life adversity is linked to distinct emotional and motivational profiles

Although participants were generally healthy, those with High ACE exposure reported significantly greater anxiety (*t*_31_ = 2.24, *p* = .032) and depressive symptoms (*z* = 2.77, *p* = .006) (**Figure 4A-B**). Depression was unrelated to state empathy or placebo responses (all *ps* > .083). In contrast, higher anxiety predicted reduced cognitive state empathy in the VR Avatar condition for both pain intensity (*r* = –0.35, *p* = .021) and pain unpleasantness (*r* = –0.38, *p* = .011), as well as in the 2D Avatar condition for pain unpleasantness (*r* = –0.35, *p* = .021), regardless of ACE exposure. No associations emerged between anxiety and placebo responses (all *ps* > .069).

**Figure 4.**
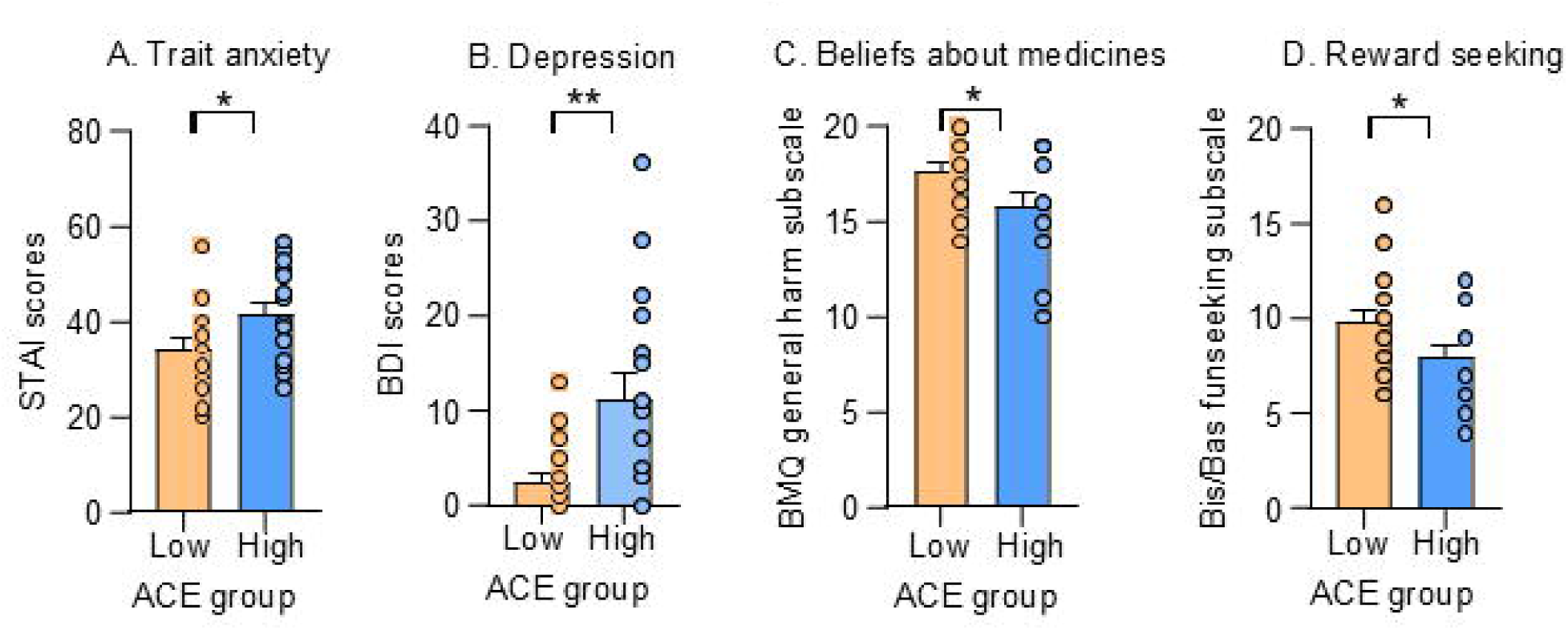
Early life adversity is linked to distinct emotional and motivational profiles. **(A-B)** Participants with higher ACE exposure reported greater anxiety and depression compared to those with low ACE exposure. **(C-D)** Participants with lower ACE exposure endorsed stronger beliefs that medicines are generally harmful and exhibited higher Fun Seeking scores, with no group differences across other BIS/BAS subscales. Error bars represent ±SEM.

Participants with Low ACE exposure endorsed stronger beliefs that medicines are generally harmful (*t*_31_ = 2.20, *p* = .036) and showed higher Fun Seeking scores (*t*_31_ = 2.14, *p* = .040) (**Figure 4 C-D**), with no group differences in General Overuse or other BIS/BAS subscales (all *ps* > .05). Neither belief nor reward-seeking traits were associated with placebo effects (all *ps* > .130). However, stronger General Harm beliefs correlated with greater cognitive state empathy for pain unpleasantness in the VR Human condition (*r* = 0.32, *p* = .027).

Together, these findings indicate that ELA is associated with heightened emotional distress and distinct motivational and belief profiles, which in turn shape social-cognitive traits such as state empathy, even if they do not directly modulate placebo responsiveness.

Mediation analyses confirmed that anxiety partially accounted for the effect of ACE exposure on cognitive state empathy, with significant indirect effects for VR Avatar pain intensity (*ab* = –0.36, 95% BCI [–0.83, –0.02]) and pain unpleasantness (*ab* = –0.36, 95% BCI [–0.76, –0.03]), and for 2D Avatar pain unpleasantness (*ab* = –0.32, 95% BCI [–0.67, –0.02]) **Figure 5**. These results suggest that ELA may blunt state empathy partly through elevated anxiety.

**Figure 5.**
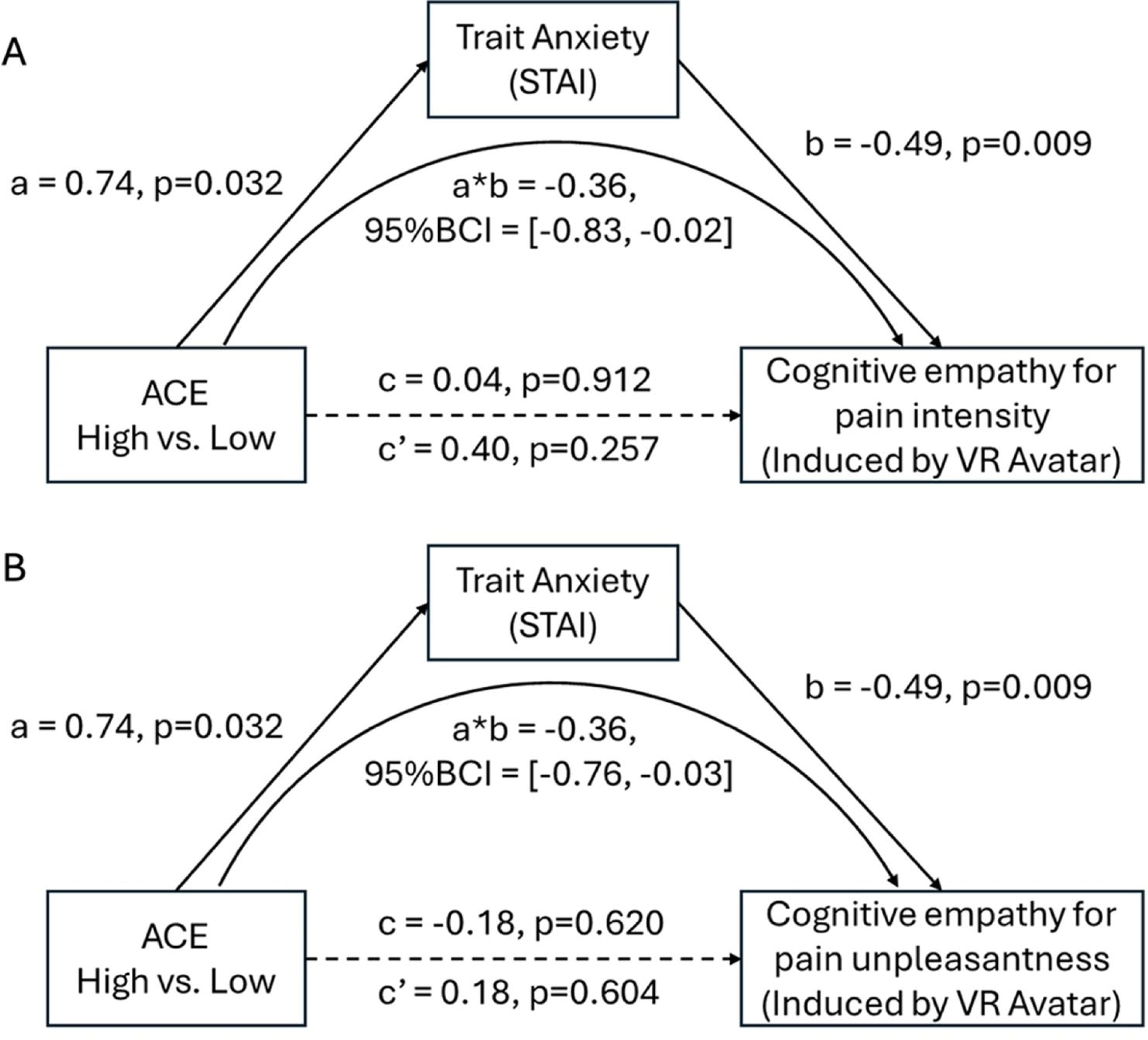
Mediation model of trait anxiety on the influences of ACE on cognitive state empathy when observing an Avatar in VR. (A). There was a partial mediation effect of anxiety on cognitive state empathy for pain intensity (sensory, other-referred). (B). There was a partial mediation effect of trait anxiety on cognitive state empathy for pain unpleasantness (emotional, other-referred).

## Discussion

This study used an observational learning procedure into immersive VR to investigate socially induced placebo analgesia and state empathy among adults stratified by low and high ACE exposure.

Contrary to our hypothesis, participants with higher ACE exposure showed stronger placebo analgesia, particularly in VR environments, whereas participants with lower ACE exposure showed context-dependent effects limited primarily to the 2D Human condition. Parallel patterns were observed for pain unpleasantness ratings, further indicating a role of ACE in learned placebo responses. The learned placebo response was unrelated to affective state empathy, which was lower in participants with high ACE exposure. Cognitive state empathy remained stable across groups. Neither cognitive nor affective state empathy mediated the observed modulation of placebo effects by ACE.

A key contribution of this work lies in demonstrating that ELA changed the direction and efficacy of socially induced placebo analgesia. There is little direct evidence of higher placebo responses in individuals with high exposure to adverse childhood events [54; 55]. Recent placebo-controlled studies examining the impact of childhood adversity on subjective drug effects in healthy adults found that greater childhood adversity was associated with dampened (not heightened) subjective responses to stimulant drugs, suggesting a blunted reward or placebo-like response rather than an enhanced one [8].

However, interventions which harness endogenous modulation — such as mindfulness therapies – enhance tolerance for cold pain and decreased cortisol levels in women with a history of childhood abuse [1]. Mindfulness interventions have also shown efficacy in reducing maladaptive and distressing behaviors in those with ACEs in multiple contexts [34; 44], suggesting prior evidence of reduced placebo responses is not reflective of impairments in endogenous pain modulatory abilities.

The observation of higher placebo effects in VR environments among individuals with high exposure to ELA is most consistent with attention or coping mechanism. VR environments are known to increase engagement compared to script-driven imagery in healthy controls [45] and particularly in individuals with heightened stress reactivity due to childhood trauma [44]. In these individuals, VR may facilitate greater attentional focus and immersive distraction [53; 60], which can enhance the perceived efficacy of placebo interventions by modulating stress and coping responses.

Specifically, individuals with high ACE exposure demonstrate increased subjective anxiety at baseline [22], suggesting that immersive environments may amplify their engagement with interventions and activate coping strategies, such as cognitive distraction or emotional regulation attempts [49]. This heightened engagement can potentiate placebo effects by leveraging attention and the individual’s coping repertoire in response to the immersive stimulus. Therefore, we speculate that the increased observationally induced placebo effects in the VR context likely reflects the interaction between VR-induced attentional capture and the coping mechanisms mobilized by individuals with a history of adversity, rather than a direct neurobiological effect of ACEs alone.

The enhanced placebo effects observed in VR environments among individuals with high exposure to ELA is most likely mediated by cognitive reappraisal [33]. Cognitive reappraisal, which involves reframing the meaning of emotionally salient stimuli, engages prefrontal cognitive control networks and is associated with greater emotional regulation and adaptive behavioral responses. Individuals who use cognitive reappraisal experience amplified benefits from VR exposure, including increased subjective vitality and reduced emotional distress [49]. Moreover, studies show that distraction-based VR interventions are equally effective regardless of coping style and avoidance does not moderate response to VR-based distraction [46].

### ELA and state empathy

Affective state empathy did not mirror the behavioral placebo outcomes in line with previous studies that demonstrated trait empathy but not state empathy [31] is associated with social observation-induced analgesia [32; 56]. High ACE exposure was associated with blunted affective state empathy across conditions, consistent with reduced emotional attunement and prior evidence linking early adversity to attenuated empathic resonance [61]. Conversely, Low ACE participants reported higher affective state empathy, suggesting that immersive VR enhances empathic engagement primarily in those without early adversity. Interestingly, cognitive state empathy remained stable across ACE groups and contexts, indicating that the ability to infer others’ mental states may be preserved after ELA. Neither affective nor cognitive state empathy mediated the relationship between ACE and placebo analgesia, implying that attentional and coping processes, rather than empathic resonance per se, may underline the observed group differences.

### A dual mechanism of socially learned placebo effects

These results might support a dual mechanism of socially learned placebo effects [2; 39]. Low ACE individuals appear to engage context-sensitive associative learning pathways that facilitate rapid expectation updating from observed relief cues. We can speculate that High ACE participants may recruit frontoparietal attentional and reappraisal networks to modulate pain perception through deliberate top-down control rather than social-affective learning. This dissociation suggests that early adversity may shift the balance between social-affective and cognitive routes to pain modulation, with immersive VR providing an alternative pathway for adaptive coping.

Overall, this is to our knowledge the first finding related to early life adversity and social observation in VR and could have important implications for therapeutic interventions. VR-based environments may offer a promising platform for enhancing engagement, promoting attentional control and reappraisal mechanisms that, in turn, facilitate symptom improvements. This is especially significant as those with high ACE scores are at an increased risk for many disorders [14; 22], which may benefit from such VR-based interventions.

## Limitations

Several limitations should be acknowledged. First, our study has a small simple size, thus causal inferences regarding mechanisms remain limited due to the cross-sectional design. Second, the assessment of ACE relied on retrospective self-report, which may be subject to recall biases. Prospective or multimodal measures of early adversity would strengthen future investigations. Third, our study did not examine how specific forms of adversity—such as sexual abuse or parental absence—differentially influence socially learned analgesia and empathy, despite evidence suggesting distinct impacts on empathic processes [17; 30]. Future studies should recruit larger or targeted samples to enable ACE type–specific analyses of placebo effects and VR efficacy. Finally, VR engagement [45] was not directly quantified beyond condition assignment. Future studies should incorporate indices of presence to delineate how immersive engagement contributes to analgesia.

## Conclusions

In conclusion, the present study suggests that high early life adversity is associated with increased sensitivity to social observation-induced placebo analgesia, especially in an immersive context. Due to the small sample size and retrospective assessment of ACE, our results should be confirmed with a larger sample and a prospective study design where ACE is assessed by independent observers.

## Declaration statements

Data Availability: All behavioral data and the VR video footage we created for this study are available upon request to the corresponding author.

## Abbreviations

BAI: Beck Anxiety Inventory
BDI: Beck Depression Inventory
BES: Basic Empathy Scale
IRI: Interpersonal Reactivity Index
STAI: State-Trait Anxiety Inventory.

## Supporting information

Supplemental FlowChart

## Data Availability

All behavioral data and the VR video footage we created for this study are available upon request to the corresponding author.

## Acknowledgements

The authors acknowledge the support of the National Center for Complementary and Integrative Health (R01-5R01AT010333, L. Colloca) and University of Maryland Baltimore Institute for Translational and Clinical Research (5TL1TR003100-05, LW). The funding source was not involved in this work. The authors would like to thank Nandini Raghuraman for helping with the procedure setup. The authors thank Drs. Yavin Shaham and Nandini Raghuraman for their support.

## Author Contributions

L.W. and L.C., conceived and designed the study. J. N. W., R. S and L.W. collected the data. J. M. H. and S. L. created the Avatar and Human demonstrator contents. G.C., B. B., A. V., S. C. helped design the study. L.W., and Y.W. performed data analysis. L.W. drafted the initial version of the manuscript. L.C edited and reviewed the final version of the manuscript. L.C. approved the final version and took responsibility for all aspects of this study.

All authors contributed to manuscript revisions and approved the final version for submission.

## References

[1] Andersen E, Geiger P, Schiller C, Bluth K, Watkins L, Zhang Y, Xia K, Tauseef H, Leserman J, Girdler S, Gaylord S. Effects of Mindfulness-Based Stress Reduction on Experimental Pain Sensitivity and Cortisol Responses in Women With Early Life Abuse: A Randomized Controlled Trial. Psychosom Med 2021;83(6):515–527.

[2] Bajcar EA, Babel P. Social Learning of Placebo Effects in Pain: A Critical Review of the Literature and a Proposed Revised Model. J Pain 2024;25(9):104585.

[3] Bajcar EA, Babel P. The role of observational learning in the formation of placebo and nocebo effects. Handb Clin Neurol 2025;213:59–69.

[4] Bajcar EA, Klosowska J, Meeuwis S, Rubanets D, Badzinska J, Babel P. Who do we learn pain from? The influence of the demonstrator’s pain assessment skills on nocebo hyperalgesia induced via observational learning. J Pain 2025;32:105420.

[5] Beck A, Steer R, Brown G. Beck Depression Inventory-II Manual, 1996.

[6] Beck AT, Epstein N, Brown G, Steer R. Beck anxiety inventory. Journal of consulting and clinical psychology 1993.

[7] Betti V, Zappasodi F, Rossini PM, Aglioti SM, Tecchio F. Synchronous with your feelings: sensorimotor {gamma} band and empathy for pain. J Neurosci 2009;29(40):12384–12392.

[8] Carlyle M, de Wit H, Leknes S. Impact of childhood adversity on acute subjective effects of stimulant and opioid drugs: Evidence from placebo-controlled studies in healthy volunteers. J Psychopharmacol 2024;38(11):986–997.

[9] Cloninger CR, Przybeck TR, Svrakic DM. The tridimensional personality questionnaire: US normative data. Psychological reports 1991;69(3):1047–1057.

[10] Colloca L, Benedetti F. Placebo analgesia induced by social observational learning. Pain 2009;144(1-2):28–34.

[11] Daskalakis NP, Bagot RC, Parker KJ, Vinkers CH, de Kloet ER. The three-hit concept of vulnerability and resilience: toward understanding adaptation to early-life adversity outcome. Psychoneuroendocrinology 2013;38(9):1858–1873.

[12] Davis MA. A multidimensional approach to individual differences in empathy. JSAS Cat Selected Docs Psychol 1980;10:85.

[13] Erdfelder E, Faul F, Buchner A. GPOWER: A general power analysis program. Behavior Research Methods, Instruments & Computers 1996;28(1):1–11.

[14] Felitti VJ, Anda RF, Nordenberg D, Williamson DF, Spitz AM, Edwards V, Koss MP, Marks JS. Relationship of childhood abuse and household dysfunction to many of the leading causes of death in adults. The Adverse Childhood Experiences (ACE) Study. Am J Prev Med 1998;14(4):245–258.

[15] Fenoglio KA, Chen Y, Baram TZ. Neuroplasticity of the hypothalamic-pituitary-adrenal axis early in life requires recurrent recruitment of stress-regulating brain regions. J Neurosci 2006;26(9):2434–2442.

[16] Fink G, Venkataramani AS, Zanolini A. Early life adversity, biological adaptation, and human capital: evidence from an interrupted malaria control program in Zambia. J Health Econ 2021;80:102532.

[17] Fourie MM, Warton FL, Derrick-Sleigh T, Codrington H, Solms M, Decety J, Stein DJ. Childhood abuse and neglect are differentially related to perceived discrimination and structural change in empathy-related circuitry. Sci Rep 2025;15(1):16361.

[18] Fruhstorfer H, Lindblom U, Schmidt WC. Method for quantitative estimation of thermal thresholds in patients. J Neurol Neurosurg Psychiatry 1976;39(11):1071–1075.

[19] Geckeler KC, Barch DM, Karcher NR. Associations between social behaviors and experiences with neural correlates of implicit emotion regulation in middle childhood. Neuropsychopharmacology 2022;47(6):1169–1179.

[20] Greenwald AG, McGhee DE, Schwartz JL. Measuring individual differences in implicit cognition: the implicit association test. J Pers Soc Psychol 1998;74(6):1464–1480.

[21] Harris PA, Taylor R, Minor BL, Elliott V, Fernandez M, O’Neal L, McLeod L, Delacqua G, Delacqua F, Kirby J. The REDCap consortium: building an international community of software platform partners. Journal of biomedical informatics 2019;95:103208.

[22] Heim C, Nemeroff CB. The role of childhood trauma in the neurobiology of mood and anxiety disorders: preclinical and clinical studies. Biol Psychiatry 2001;49(12):1023–1039.

[23] Honzel E, Murthi S, Brawn-Cinani B, Colloca G, Kier C, Varshney A, Colloca L. Virtual reality, music, and pain: developing the premise for an interdisciplinary approach to pain management. Pain 2019;160(9):1909–1919.

[24] Jiang L, Xue L, Juruena MF. The impact of early life stress on the hypothalamic-pituitary-adrenal axis in unipolar major depression: A systematic review. Psychoneuroendocrinology 2025;181:107607.

[25] Jolliffe D, Farrington DP. Development and validation of the Basic Empathy Scale. Journal of adolescence 2006;29(4):589–611.

[26] Malave L, van Dijk MT, Anacker C. Early life adversity shapes neural circuit function during sensitive postnatal developmental periods. Transl Psychiatry 2022;12(1):306.

[27] Mathur A, Graham-Engeland JE, Slavish DC, Smyth JM, Lipton RB, Katz MJ, Sliwinski MJ. Recalled early life adversity and pain: the role of mood, sleep, optimism, and control. J Behav Med 2018;41(4):504–515.

[28] McEwen CA, Gregerson SF. A Critical Assessment of the Adverse Childhood Experiences Study at 20 Years. Am J Prev Med 2019;56(6):790–794.

[29] McKibben LA, Woolard A, McLean SA, Zhao Y, Verma T, Mickelson J, Lu H, Lobo J, House SL, Beaudoin FL, An X, Stevens JS, Neylan TC, Jovanovic T, Germine LT, Rauch SL, Haran JP, Storrow AB, Lewandowski C, Hendry PL, Sheikh S, Jones CW, Punches BE, Hudak LA, Pascual JL, Seamon MJ, Pearson C, Peak DA, Merchant RC, Domeier RM, Rathlev NK, O’Neil BJ, Sanchez LD, Bruce SE, Sheridan JF, Kessler RC, Koenen KC, Ressler KJ, Linnstaedt SD. Early life adversity increases risk for chronic post-traumatic pain, data from humans and rodents. Pain 2025;166(9):2103–2115.

[30] McLaughlin KA, Sheridan MA, Humphreys KL, Belsky J, Ellis BJ. The Value of Dimensional Models of Early Experience: Thinking Clearly About Concepts and Categories. Perspect Psychol Sci 2021;16(6):1463–1472.

[31] Meeuwis SH, Klosowska J, Budzisz A, Jankowska A, Rubanets D, Badzinska J, Bajcar EA, Babel P. I do not feel your pain: Exploring the impact of state empathy on placebo and nocebo effects evoked by observational learning. J Pain 2025;35:105526.

[32] Meeuwis SH, Wasylewski MT, Bajcar EA, Bieniek H, Adamczyk WM, Honcharova S, Di Nardo M, Mazzoni G, Babel P. Learning pain from others: a systematic review and meta-analysis of studies on placebo hypoalgesia and nocebo hyperalgesia induced by observational learning. Pain 2023;164(11):2383–2396.

[33] Moseley GL, Butler DS. Fifteen Years of Explaining Pain: The Past, Present, and Future. J Pain 2015;16(9):807–813.

[34] Moyes E, Nutman G, Mirman JH. The Efficacy of Targeted Mindfulness-Based Interventions for Improving Mental Health and Cognition Among Youth and Adults with ACE Histories: A Systematic Mixed Studies Review. J Child Adolesc Trauma 2022;15(4):1165–1177.

[35] Peltonen K, Tammilehto J, Flykt M, Vanska M, Kuppens P, Bosmans G, Lindblom J. Adverse childhood experiences and emotion dynamics in daily life: a two sample study. Cogn Emot 2025;39(6):1250–1270.

[36] Persky S, Colloca L. Medical Extended Reality Trials: Building Robust Comparators, Controls, and Sham. J Med Internet Res 2023;25:e45821.

[37] Rabin J, Lawlace M, Zhen-Duan J, Nunez M, Jacquez F. A social interaction learning model approach to understand adverse childhood experiences and drug use among Latinx youth. J Ethn Subst Abuse 2023;22(3):644–658.

[38] Raghuraman N, Wang Y, Schenk LA, Furman AJ, Tricou C, Seminowicz DA, Colloca L. Neural and behavioral changes driven by observationally-induced hypoalgesia. Sci Rep 2019;9(1):19760.

[39] Raghuraman N, White JN, Watson L, Bellei-Rodriguez CE, Shafir R, Wang Y, Colloca L. Neuropsychological mechanisms of observational learning in human placebo effects. Psychopharmacology (Berl) 2025;242(5):889–900.

[40] Rouch I, Strippoli MF, Dorey JM, Laurent B, Ranjbar S, Marques-Vidal PM, Berna C, Suter M, Vaucher J, von Gunten A, Preisig M. Early-life adversity predicting the incidence of multisite chronic pain in the general population. Eur Psychiatry 2024;67(1):e67.

[41] Schellhaas S, Schmahl C, Bublatzky F. Social threat and safety learning in individuals with adverse childhood experiences: electrocortical evidence on face processing, recognition, and working memory. Eur J Psychotraumatol 2022;13(2):2135195.

[42] Schenk LA, Colloca L. The neural processes of acquiring placebo effects through observation. Neuroimage 2020;209:116510.

[43] Schenk LA, Krimmel SR, Colloca L. Observe to get pain relief: current evidence and potential mechanisms of socially learned pain modulation. Pain 2017;158(11):2077–2081.

[44] Schweizer T, Renner F, Sun D, Kleim B, Holmes EA, Tuschen-Caffier B. Psychophysiological reactivity, coping behaviour and intrusive memories upon multisensory Virtual Reality and Script-Driven Imagery analogue trauma: A randomised controlled crossover study. J Anxiety Disord 2018;59:42–52.

[45] Shafir R, Watson L, Felix RB, Muhammed S, Fisher JP, Hu P, Wang Y, Colloca L. Factors influencing the hypoalgesic effects of virtual reality. Pain 2025;166(8):1836–1846.

[46] Sil S, Dahlquist LM, Thompson C, Hahn A, Herbert L, Wohlheiter K, Horn S. The effects of coping style on virtual reality enhanced videogame distraction in children undergoing cold pressor pain. J Behav Med 2014;37(1):156–165.

[47] Spielberger CD, Gonzalez-Reigosa F, Martinez-Urrutia A, Natalicio LF, Natalicio DS. The state-trait anxiety inventory. Revista Interamericana de Psicologia/Interamerican journal of psychology 1971;5(3 & 4).

[48] Tan MCC, Chye SYL, Teng KSM. “In the shoes of another”: immersive technology for social and emotional learning. Educ Inf Technol (Dordr) 2022;27(6):8165–8188.

[49] Theodorou A, Spano G, Bratman GN, Monneron K, Sanesi G, Carrus G, Imperatori C, Panno A. Emotion regulation and virtual nature: cognitive reappraisal as an individual-level moderator for impacts on subjective vitality. Sci Rep 2023;13(1):5028.

[50] Thomas PA, Goodin BR, Meints SM, Owens MA, Wiggins AM, Quinn T, Long L, Aroke EN, Morris MC, Sorge RE, Overstreet DS. Adverse Childhood Experiences and Chronic Low Back Pain in Adulthood: The Role of Emotion Regulation. J Pain 2024;25(9):104551.

[51] Thomas PA, Van Ditta P, Stocking SQ, Webb C, Meints SM, Owens MA, Quinn T, Aroke EN, Morris MC, Sorge RE. The effects of neighborhood disadvantage and adverse childhood experiences on conditioned pain modulation in adults with chronic low back pain. The Journal of Pain 2025;26:104706.

[52] Uchida S, Hara K, Kobayashi A, Funato H, Hobara T, Otsuki K, Yamagata H, McEwen BS, Watanabe Y. Early life stress enhances behavioral vulnerability to stress through the activation of REST4-mediated gene transcription in the medial prefrontal cortex of rodents. J Neurosci 2010;30(45):15007–15018.

[53] Veling W, Counotte J, Pot-Kolder R, van Os J, van der Gaag M. Childhood trauma, psychosis liability and social stress reactivity: a virtual reality study. Psychol Med 2016;46(16):3339–3348.

[54] Waltz JA, Pujji SD, Colloca L. Placebo and nocebo phenomena in schizophrenia spectrum disorders: a narrative review on current knowledge and potential future directions. Psychol Med 2025;55:e199.

[55] Weimer K, Colloca L, Enck P. Placebo eff ects in psychiatry: mediators and moderators. Lancet Psychiatry 2015;2(3):246–257.

[56] White JN, Watson L, Wang Y, Colloca G, Heagerty JM, Li S, Brawn B, Varshney A, Shafir R, Belleï-Rodriguez C, Colloca L. Context-Dependent Placebo Hypoalgesia Through Observational Learning: The Role of Empathy in Immersive and Non-Immersive Environments. npj Digital Medicine under revision.

[57] Who. Adverse Childhood Experiences International Questionnaire (ACE-IQ), 2018.

[58] Wu CM, Deffner D, Kahl B, Meder B, Ho MK, Kurvers R. Adaptive mechanisms of social and asocial learning in immersive collective foraging. Nat Commun 2025;16(1):3539.

[59] Yam KY, Ruigrok SR, Ziko I, De Luca SN, Lucassen PJ, Spencer SJ, Korosi A. Ghrelin and hypothalamic NPY/AgRP expression in mice are affected by chronic early-life stress exposure in a sex-specific manner. Psychoneuroendocrinology 2017;86:73–77.

[60] Yan S, Zhong S, Lyu S, Lai S, Zhang Y, Luo Y, Ran H, Duan M, Jia Y. A randomized trial of virtual reality eye movement desensitization and reprocessing therapy for major depressive disorder with childhood trauma: A 3-month follow-up study. Psychol Trauma 2025;17(5):1127–1139.

[61] Zhang H, Gao X, Liang Y, Yao Q, Wei Q. Does Child Maltreatment Reduce or Increase Empathy? A Systematic Review and Meta-analysis. Trauma Violence Abuse 2024;25(1):166–182.

[62] Zhang X, Merrin GJ, Slavich GM. Adverse childhood experiences (ACEs) and emotion dysregulation phenotypes: An intersectional analysis of race/ethnicity and gender in a nationally representative U.S. sample. Child Abuse Negl 2024;158:107129.

[63] Zimmer C, Spencer KA. Modifications of glucocorticoid receptors mRNA expression in the hypothalamic-pituitary-adrenal axis in response to early-life stress in female Japanese quail. J Neuroendocrinol 2014;26(12):853–860.

