## Supplemental FlowChart for "Effects of childhood adversity on socially learned placebo analgesia in virtual reality: A cross-sectional study"

**Flow chart**:

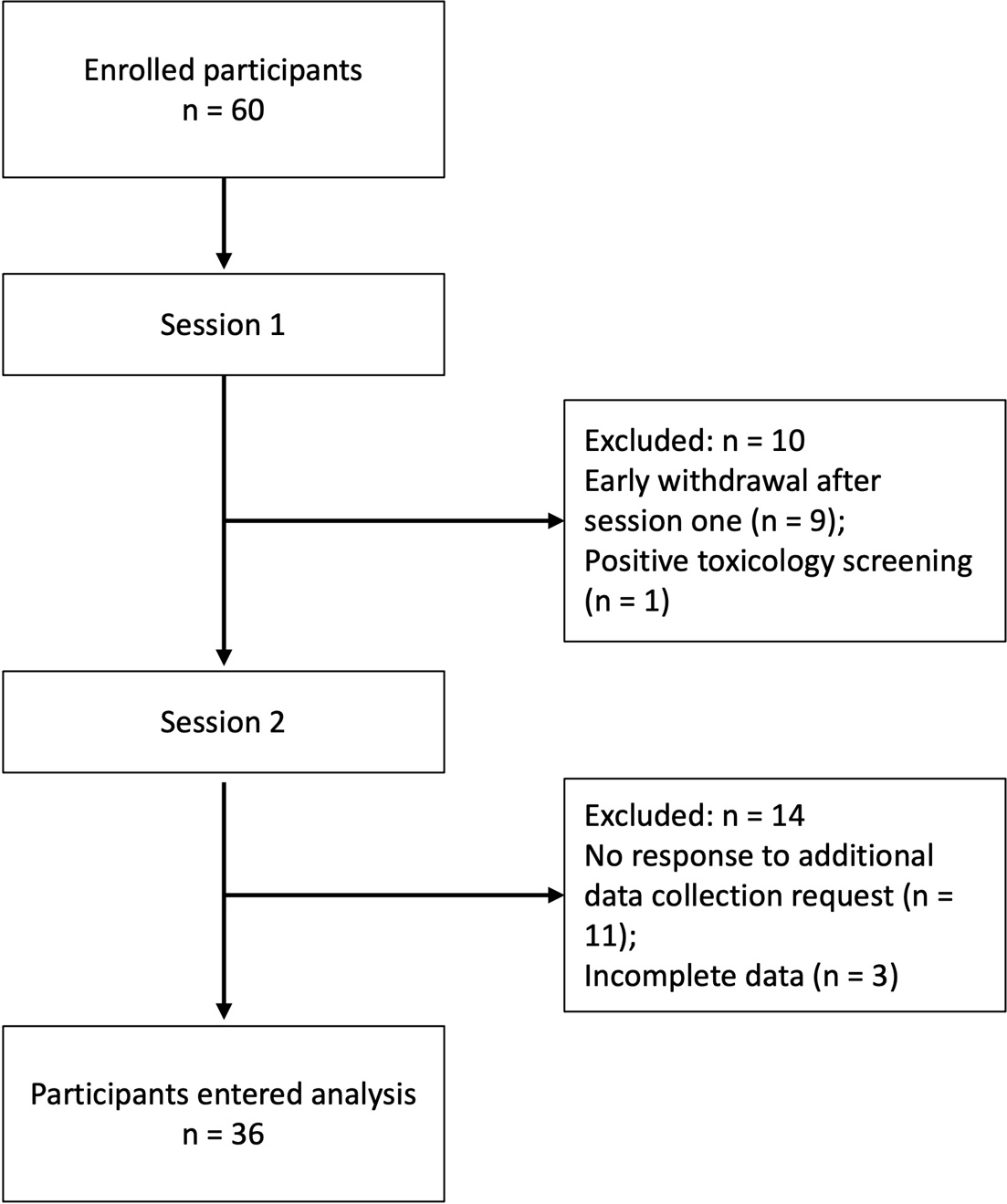

Twenty-four participants were excluded from protocol for early withdrawal after session one (n = 9); a positive toxicology screening (n = 1); incomplete data (n = 3); and lack or response to request for additional data collection (n = 11).
